# Deep learning models for COVID-19 infected area segmentation in CT images

**DOI:** 10.1101/2020.05.08.20094664

**Authors:** Athanasios Voulodimos, Eftychios Protopapadakis, Iason Katsamenis, Anastasios Doulamis, Nikolaos Doulamis

## Abstract

Recent studies indicated that detecting radiographic patterns on CT chest scans can yield high sensitivity and specificity for COVID-19 detection. In this work, we scrutinize the effectiveness of deep learning models for semantic segmentation of pneumonia infected area segmentation in CT images for the detection of COVID-19. We explore the efficacy of U-Nets and Fully Convolutional Neural Networks in this task using real-world CT data from COVID-19 patients. The results indicate that Fully Convolutional Neural Networks are capable of accurate segmentation despite the class imbalance on the dataset and the man-made annotation errors on the boundaries of symptom manifestation areas, and can be a promising method for further analysis of COVID-19 induced pneumonia symptoms in CT images.

**Impact Statement:** Fully Convolutional Neural Networks appear to be an accurate segmentation method in CT scans for COVID-19 pneumonia and could assist in the detection as a fast and cost-effective option.

## I. Introduction

THE novel coronavirus 2019-nCoV first transmitted to humans in December 2019, resulting in a pandemic outbreak the following months. The disease, known as COVID-19 [1] caused or is expected to cause significant short-term and long-term societal and economic impacts [2], resulting in more than 260,000 deaths up to 7^th^ of May 2020 [3].

A further insight on the findings so far indicate that COVID-19 affects multiple organs in the human body, including heart and blood vessels, kidneys, gut, and brain. The virus enters the cells by binding to surface receptors angiotensin-converting enzyme 2 or ACE2. This receptor can be found on alveoli, i.e. tiny air sacs in human lungs. Thus, lungs become the ground zero for the virus affection [4].

In this context, CT scanning could be a promising and efficient alternative or auxiliary tool for the detection and control of COVID-19 disease, compared to other types of tests. For example, a test based on reverse transcription polymerase chain reaction (RTPCR), takes 4 to 6 hours, assuming that the required resources are available.

CT scan analysis can be interpreted as an image analysis problem, which can be addressed as: a) classification, b) object detection, and c) semantic segmentation problem. The first approach, i.e. CT scan classification provides a binary outcome of the form 0 or 1, which indicates if the patient has COVID-19. The second approach, in case of a positive detection, provides bounding boxes, indicated the symptomatic areas. The third case involves the pixel level detection of the symptomatic areas, in each of the CT scan slices. In this paper, we propose a deep learning semantic segmentation approach for the annotation of symptomatic lung areas, for COVID-19 patients.

### A. Related Work

CT abnormalities related to COVID-19 patients, are a common case and are reported and used by the doctors in multiple studies [5]–[7]. There are two important outcomes from these studies: a) there are clear patterns indicating viral infections, even at early stages [6], [7], and b) CT abnormalities diagnostic of viral pneumonia can be available before a positive laboratory test in almost 70% of the cases [5]. Hence, CT investigation appears a promising candidate for an early detection of COVID-19 infections.

Research outcomes on COVID-19 confirmed cases, indicated that CT abnormalities, before the appearance of clinical symptoms, may occur [8]. Asymptomatic patients typically have abnormal chest CT, which are consistent with viral pneumonia. On the one hand, typical patterns may refer to unilateral, multifocal and peripherally based ground glass opacities [GGO]. On the other hand, interlobular septal thickening, thickening of the adjacent pleura, nodules, round cystic changes, bronchiectasis, pleural effusion, and lymphadenopathy were rarely observed in the asymptomatic group, but appear in symptomatic cases.

The adaptation of any visual detection approach should emphasize on the identification of predominant patterns of lung abnormalities like GGOs, crazy-paving pattern, consolidation, and linear opacities. Yet, the appearance rates and the density varied greatly depending on the stage of the disease, expecting a maximum manifestation after 9 days from the onset of the initial symptoms [6].

Deep learning approaches over various types of images consist a common approach for identification, detection or segmentation in medical imaging [9]. In this context, several approaches have already started being investigated by researchers to assist medical professionals in COVID-19 detection.

A first approach was the classification of multiple CT slices using a convolutional neural network variation [10]. The adopted methodology is capable to identify a viral infection with an ROCAUC score of 0.95 (score of 1 indicates a perfect classifier). However, despite the high detection rates, authors indicate that is extremely difficult to distinguish between different types of viral pneumonia based solely on CT analysis.

CNN variations for the distinction of Coronavirus vs Non-coronavirus cases has been proposed by [11]. The specific approach allows for the distinction between COVID-19, other types of viral infections and non-infection cases. Results indicate that there are adequate detection rates and a higher detection rate than RT-PCR testing.

A multistage approach involving segmentation and the classification between COVID-19 and other viral infection has been proposed in the work of [12], allowing for advanced disease progression monitoring. At first, a segmentation approach, i.e. U-Net, focus on the lungs regions, by removing image portions that are not relevant for the detection. Then, a pretrained Resnet-50 network is modified to handle the classification problem: COVID-19 or other cases.

Volumetric Medical Image segmentation networks, known as V-nets [13], were also utilized. The work of [14], used a V-net to segment all the slices of a given MRI, at once. Quantitative evaluation results indicate that automatic infection region delineation can be feasible and effective.

An object detection approach, i.e. denoting the areas of interest using bounding boxes was, also, considered [15]. The detection of symptomatic lung areas has been achieved by employing a VGG architecture [16] variation. Proposed approach can classify COVID-19 cases from community acquired pneumonia (CAP) and non-pneumonia (NP).

### B. Our contribution

In this paper, we propose a deep learning framework for the identification of areas with COVID-19 symptoms in CT scans. Compared to other approaches, we adopt a light U-Net model from scratch, which can be trained and operate on ordinary PC without GPU utilization, requires a limited annotated dataset for training and validations, and can handle the class imbalance problem.

## II. Materials and Methods

Identification of COVID-19 symptoms on CT images could be seen as a binary classification approach; the negative class consists of regions without COVID-19 induced symptoms, e.g. swelling, lesions and other types, described in section I.A., and the positive class includes areas depicting symptoms manifestation related to COVID-19.

Such semantic segmentation tasks can be implemented in a two-step process: (i) feature extraction over an image patch and (ii) a training process, using annotated datasets. In such a scenario, each pixel is described by feature values, extracted locally, over a, typically, small area, denoted as “patch”. Deep learning approaches do both steps for a given set of data. The main question, thus, involves the type of deep learning approach: traditional CNNs over image patches [9] or FCNs over the entire image.

In the former case, a classifier is fed with these feature values and produces an outcome, which classifies the pixel at the center as positive or negative detection. As such, for any CT slice (image) of size 630 × 630 pixels, and given a patch size of 11 × 11 pixels, we should annotate (630 − 5 − 5) × (630 − 5 − 5) = 384,400 overlapping image patches. Deep learning feature extraction has been the common case approach; experimental results indicate the benefits over traditional, hand-made, feature extraction processes. In such case, a CNN classifier could annotate the image within a few seconds time frame [17]. The advantages of such a technique are the high accuracy rate and the flexibility in handling unbalanced data sets.

The latter case involves the utilization of the entire image and the annotation in one pass. Towards that direction, the fully convolutional neural networks techniques were considered and implemented. The main advantages of such processes are described in the next section.

### A. Employed deep learning techniques

There are various levels of granularity in image understanding, starting from a coarse-grained down to a more fine-grained comprehension. The first step is the classification. In this case, we just indicate if an image depicts a COVID infection or not. The second step includes localization, where along with the discrete label, i.e. COVID-19 or not, we also expect a bounding box, indicating the area of interest. That way, the model assists the experts by narrowing the time they have to spend on scans.

However, for many applications, bounding boxes do not suffice, e.g. precise tumor detection. In such cases, we need the information on a pixel-level basis, i.e. highly detailed results. This is the goal of semantic image segmentation algorithms. In this case, we try to label each pixel of an image with a corresponding class of what is being represented. Semantic segmentation comes with specific limitations in the form of time constraints, limited hardware access, and low false-negative detection thresholds.

In this study, we handle the semantic segmentation problem for COVID-19 infection-induced symptoms in the lung areas, given as inputs CT scans. Fig. 1 demonstrates the proposed approach outcomes compared to other segmentation approaches or experts’ annotated images.

**Fig. 1.**
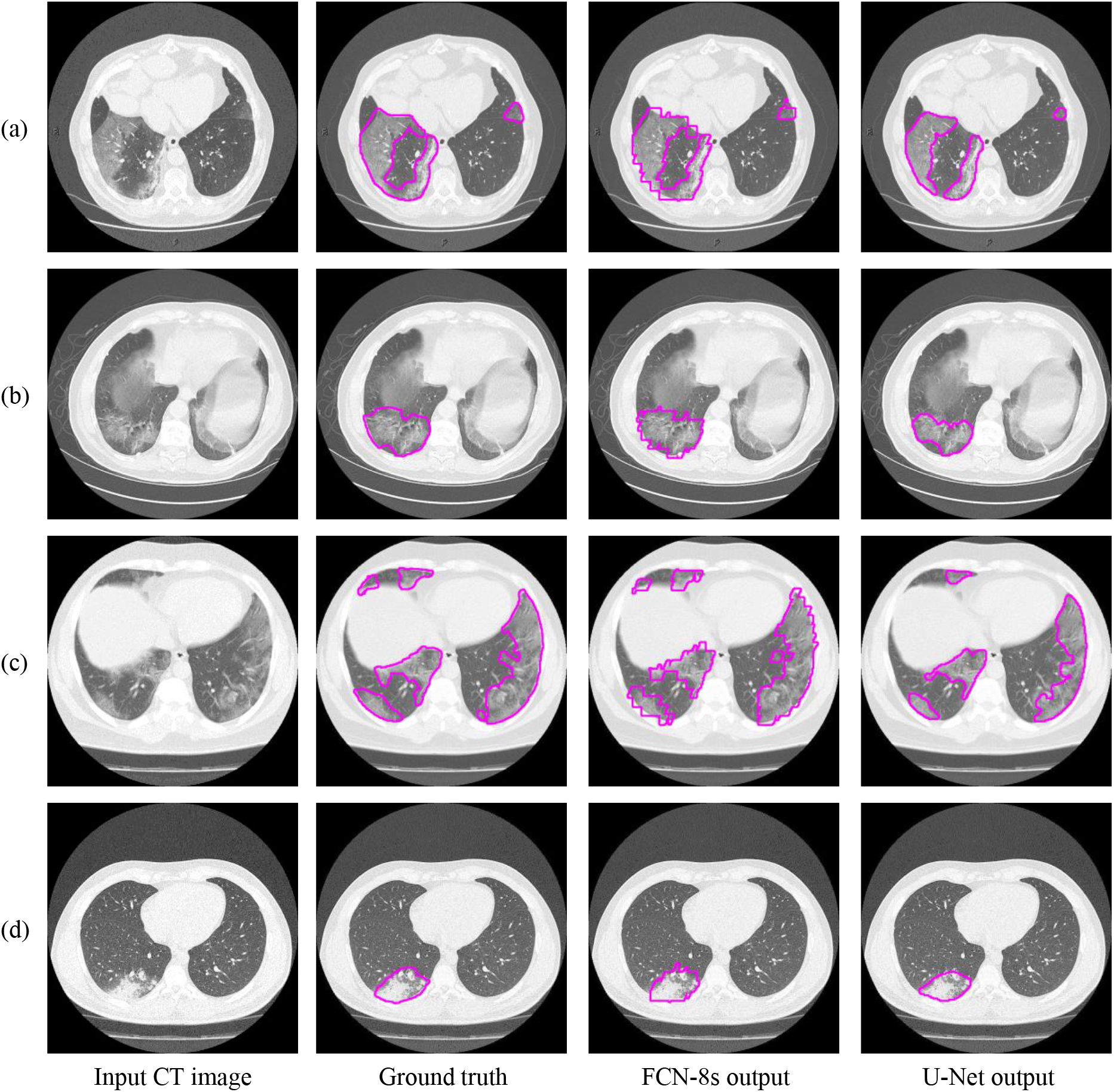
Semantic segmentation results comparison among deep learning models’ outputs and experts’ annotations.

#### 1) Fully convolutional neural networks

Fully Convolutional Networks (FCNs), as the name suggests, are built using locally connected layers, such as convolution, pooling and upsampling [18]. Note that no dense layer is used in this kind of architecture. This reduces the number of parameters and computation time [19], [20]. Their topology contains 2 parts: (i) downsampling path, which is responsible for capturing semantic/contextual information and (ii) upsampling path, responsible for recovering spatial information. Any disadvantages related to information loss, due to pooling or downsampling layers, can be mitigated using an operation called skip connection, which bypasses at least one layer.

#### 2) U-Nets

U-Net is another variation based on the CNNs, designed and applied in 2015 to process biomedical images [21], [22]. As a general convolutional neural network focuses its task on image classification, where input is an image and output is one label, but in biomedical cases, it requires us not only to distinguish whether there is a disease, but also to localize the area of abnormality.

The U-Net is built upon the Fully Convolutional Network and modified in a way that it yields better segmentation in medical imaging. To that extent, the architecture contains two paths. The first path is the contraction path (also known as the encoder) which is used to capture the context in the image. The encoder is just a traditional stack of convolutional and max-pooling layers. The second path is the symmetric expanding path (also known as the decoder) which is used to enable precise localization using transposed convolutions. Contracting and expanding paths are connected using a bottleneck, built from simply 2 convolutional layers (with batch normalization), with dropout.

Compared to known FCN approaches, e.g. FCN-8s [18], the two main differences are the (i) symmetry and (ii) connection skipping between paths. U-Net is symmetric. Furthermore, the skip connections between the downsampling path and the upsampling path apply a concatenation operator instead of a sum. These skip connections intend to provide local information to the global information while upsampling. Given the model’s symmetry, the network has a large number of feature maps in the upsampling path, which allows transferring information.

Fig. 2 provides further insights regarding the models’ annotations, in terms of accuracy and edge smoothness. Given a CT scan slice, the FCN-8 model tends to produce more coarse boundaries. On the other hand, U-Net provide smoother regions, slightly smaller that the original annotated area. Both models are capable to localize well, for the majority of symptomatic regions.

**Fig. 2.**
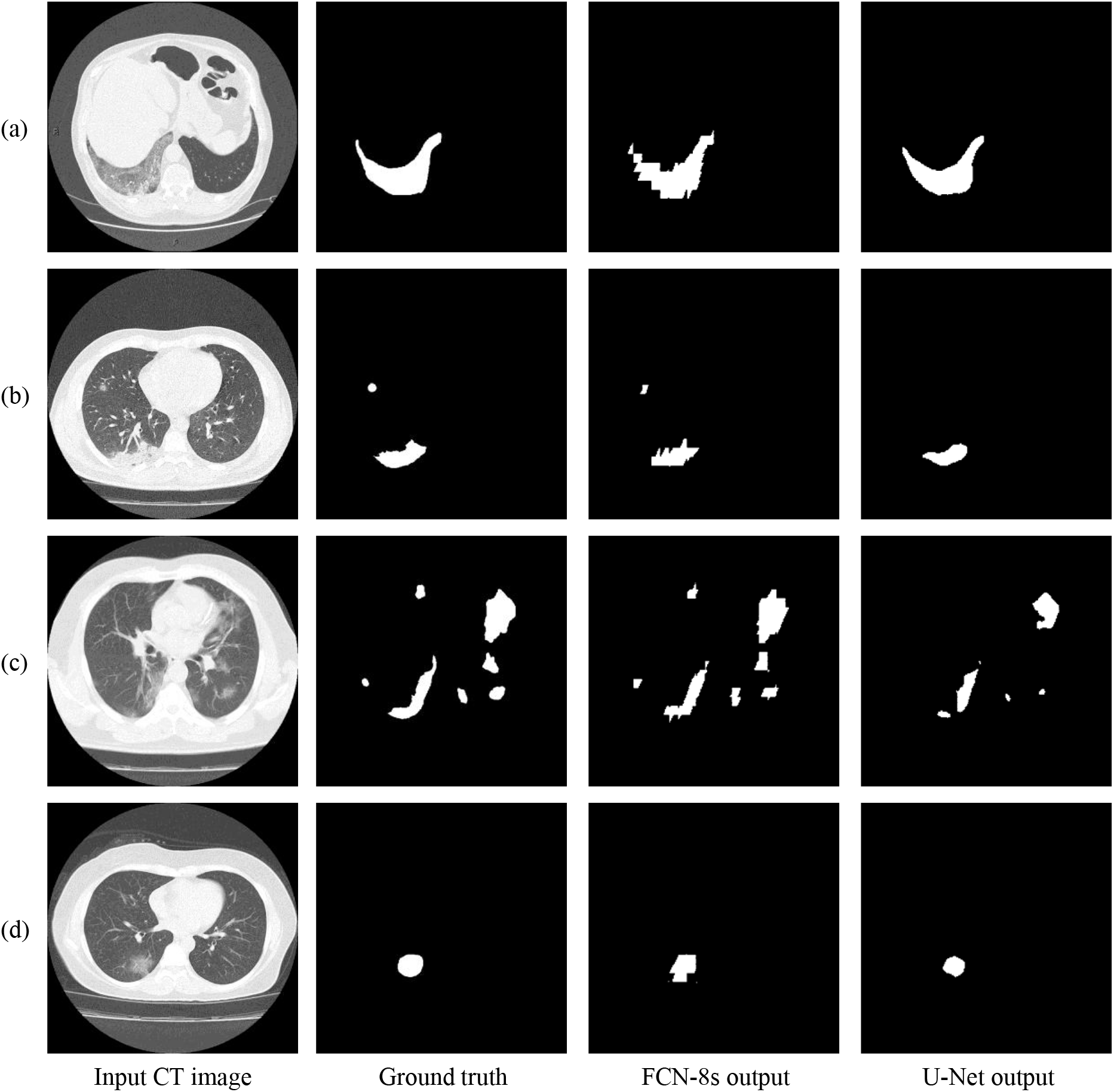
Visual comparison of the deep models’ outputs. The leftmost column is the original CT scan image, whereas the second column illustrates the corresponding segmentation for COVID-19 symptomatic areas. The last two columns depict the generated semantic segmented area.

## III. Results and Discussion

All models were developed in Python, using Keras and TensorFlow libraries. The deep models were trained using an NVIDIA Tesla P4 GPU, provided by Google Colab. For the evaluation process we conducted tests on a typical PC with 8 CPU cores (AMD FX-8320 @ 3.5 GHz) and 8GB RAM. Fig. 3 describes the adopted topology for the proposed U-Net architecture. The final U-Net model required less than 3MB of storage space.

**Fig. 3.**
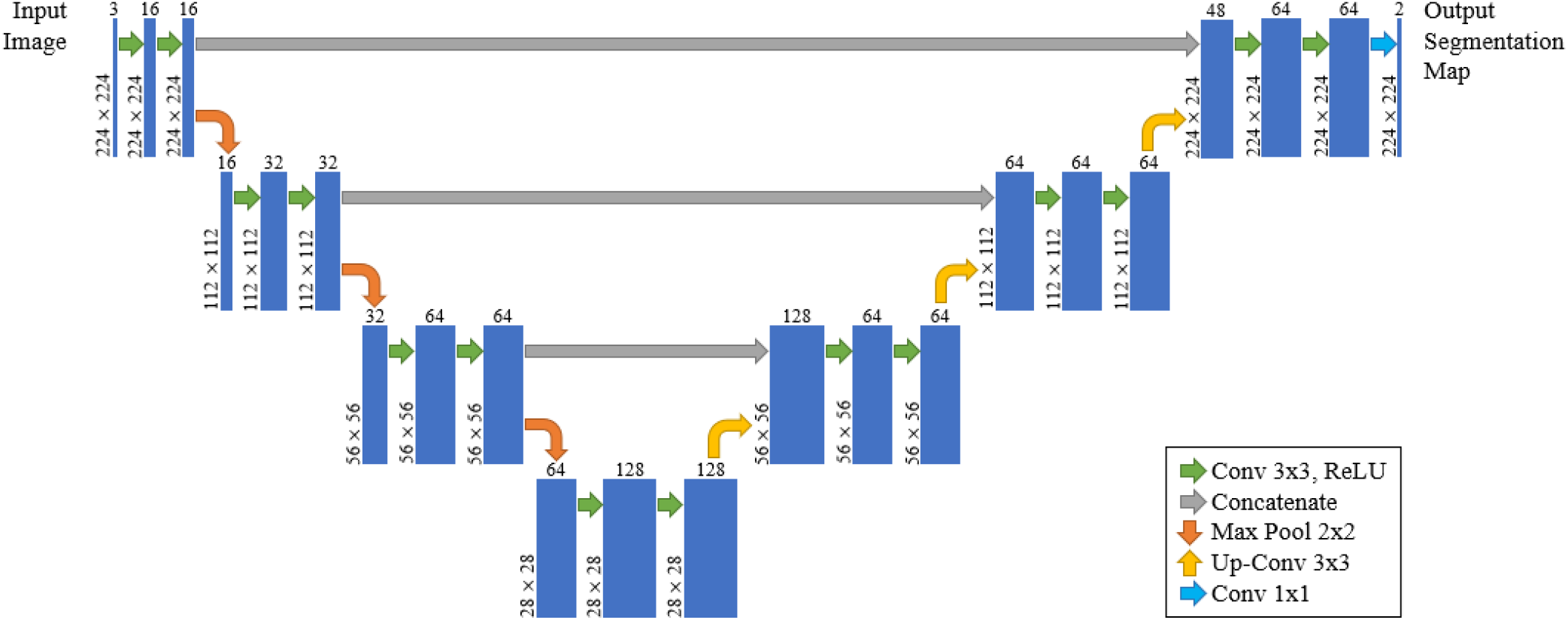
The proposed U-Net architecture.

### A. Dataset description

This dataset was collected from Radiopaedia [23] and manually annotated in the work of [24], [25]. All images are CT scans of lungs, with dimensions of 630 × 630 pixels, and were labeled, segmented, and verified by radiologist experts. More specifically, it consists of 10 axial volumetric CT scans of confirmed COVID-19 pneumonia patients. It is noted that the dataset consists of CT volumes providing a total of 939 cross-sectional images, both positive and negative. In particular, 447 slices have been labeled as negative and 492 as positive and then segmented by radiologist experts. From the whole number of CT images in the dataset, about 85% is used for training and validation of the deep learning models, while the rest 15% is used for testing. Among the training data, about 90% of them were used for training and the remaining 10% for validation.

### B. Implementation and limitations of mitigation strategies

Prior to any implementation approach, we should consider the limitations of the problem at hand. In deep learning approaches, there are two main concerns: (i) data availability and (ii) data imbalance, which both impact the classification model selection and topology’s complexity.

The first step was a training data balancing strategy, involving under-sampling of the majority class [26]. At first glance, approximately 400 images contain no positive annotations. These were excluded from the training set. The remaining 300, approximately, images had various ratios ranging from 0.1% to 20% of positive annotations to image total pixels.

Man-made annotations are prone to errors [27]. It is extremely difficult, rather impossible for most cases, to be able to distinguish if a specific pixel, on a boundary area, between two classes, corresponds to either of them. Towards that direction, we could utilize the networks’ capabilities to generalize and handling the noise, given that the wrong annotations are limited.

Other approaches considered where the implementation of different performance metrics during the training process and building models of limited complexity.

### C. Experimental results

Experimental results consider both the detection capabilities, employing multiple classification related performance metrics and the computational average time, required by a trained model to fully annotate a CT slice. Fig. 4 provides the average execution times per image, which range between 0.01 to 0.018 seconds.

**Fig. 4.**
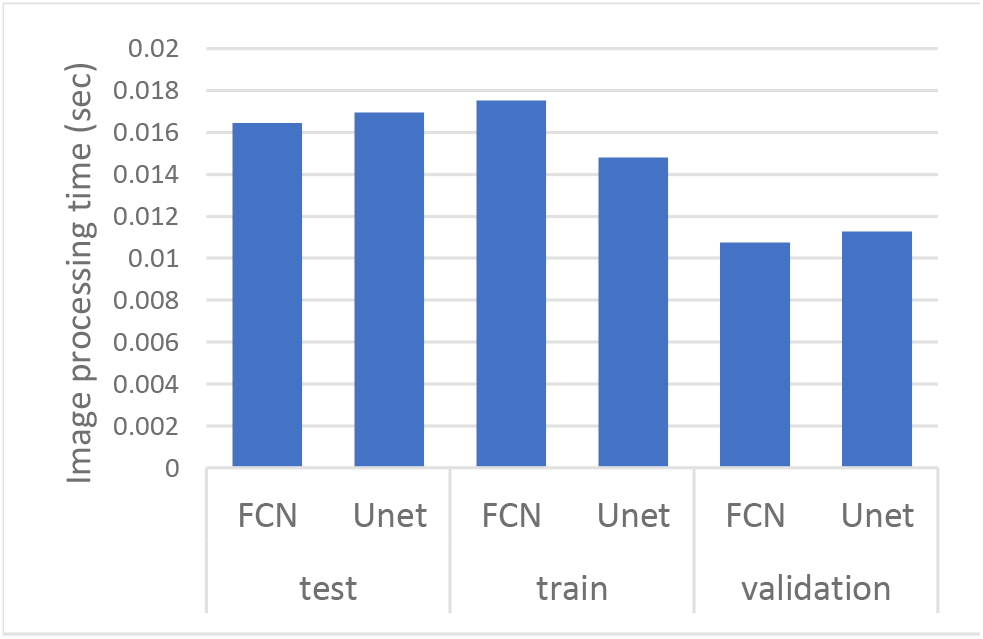
Average processing time per image.

Similarly to the evaluation strategies adopted in other classification related problems, four performance metrics were considered: a) precision, which calculates how many correct positive predictions we have, b) recall, which indicates the fraction of the positive samples that are successfully retrieved, c) accuracy, which is the percentage of correct classification for both, positive and negative, classes, and d) F1-score, which is the weighted harmonic mean of precision and recall.

Fig. 5 illustrates the performance scores in terms of precision and recall, for the segmentation approaches in train, validation and test data sets. In this case, we mainly focus on recall. Recall indicates the model’s capability to identify the case; i.e. if a CT slice image has COVID symptomatic areas the model will indicate these areas even if precision (on the positive class) is limited. Results indicate two important aspects: significant recall variation scores between train and test sets and b) the better generalization capabilities of U-Net despite the lower performance during training and validation.

**Fig. 5.**
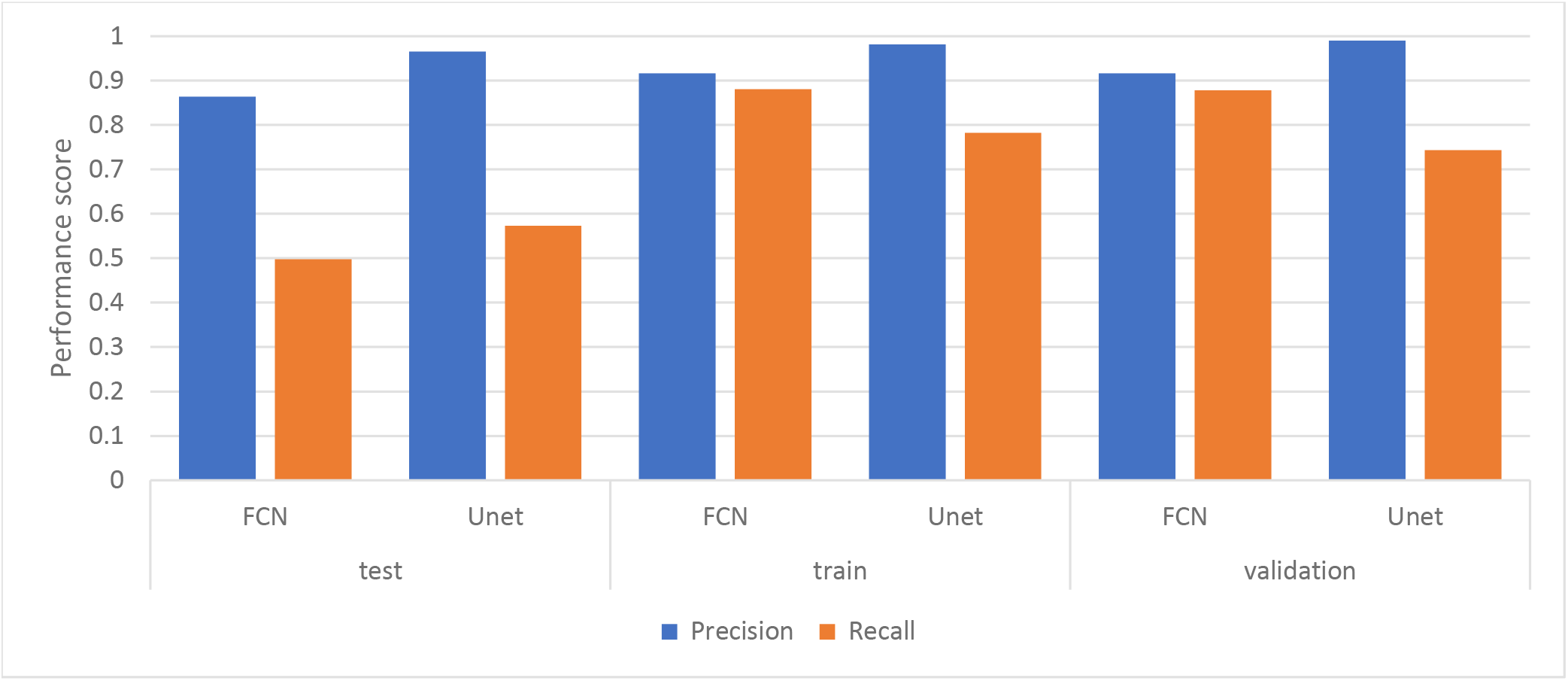
Performance results, in terms of precision and recall scores, for the segmentation approaches in train, validation and test data sets.

Fig. 6 displays the performance scores in terms of accuracy and F1 scores, for the segmentation approaches in train, validation and test datasets. The difference between accuracy and F1 score can be put down to the class imbalance. Indeed, the majority class, i.e. no detections, is almost always correctly identified. The false-negative detections can be spotted on the boundaries in images where COVID symptoms manifestation becomes apparent. Yet, the F1 score is relatively high, indicating that the minority class, i.e. COVID symptomatic areas, can be identified.

**Fig. 6.**
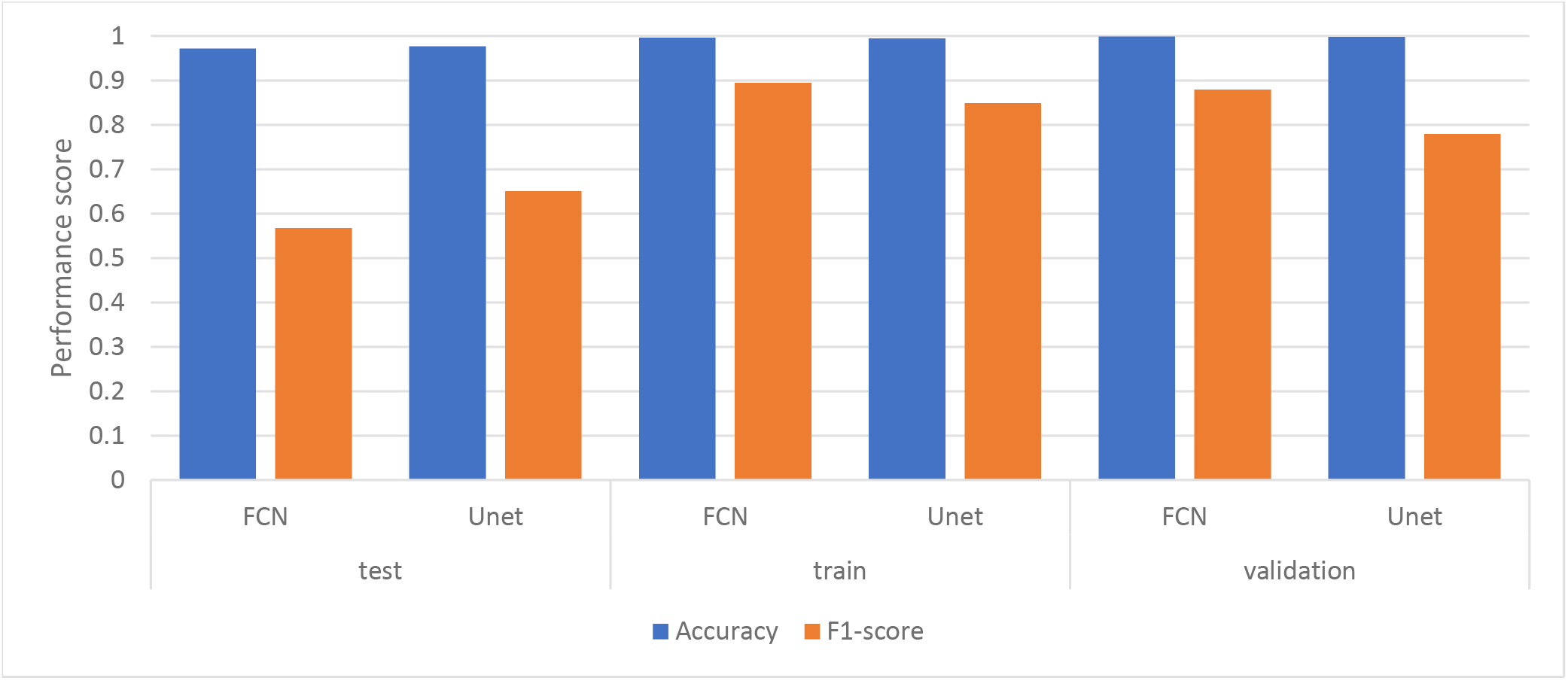
Performance results, in terms of accuracy and F1 scores, for the segmentation approaches in train, validation and test data set.

Finally, Fig. 7 provides an indication of low detection rates cases. Detection failures may include partial area annotation or non-annotation at all, despite the appearance of symptoms in the CT slice. However, CT scans have consecutive slices; even if the detection fails for the current slice, it is highly likely that it will succeed in the next ones, thus providing a potentially valuable aid to medical professionals.

**Fig. 7.**
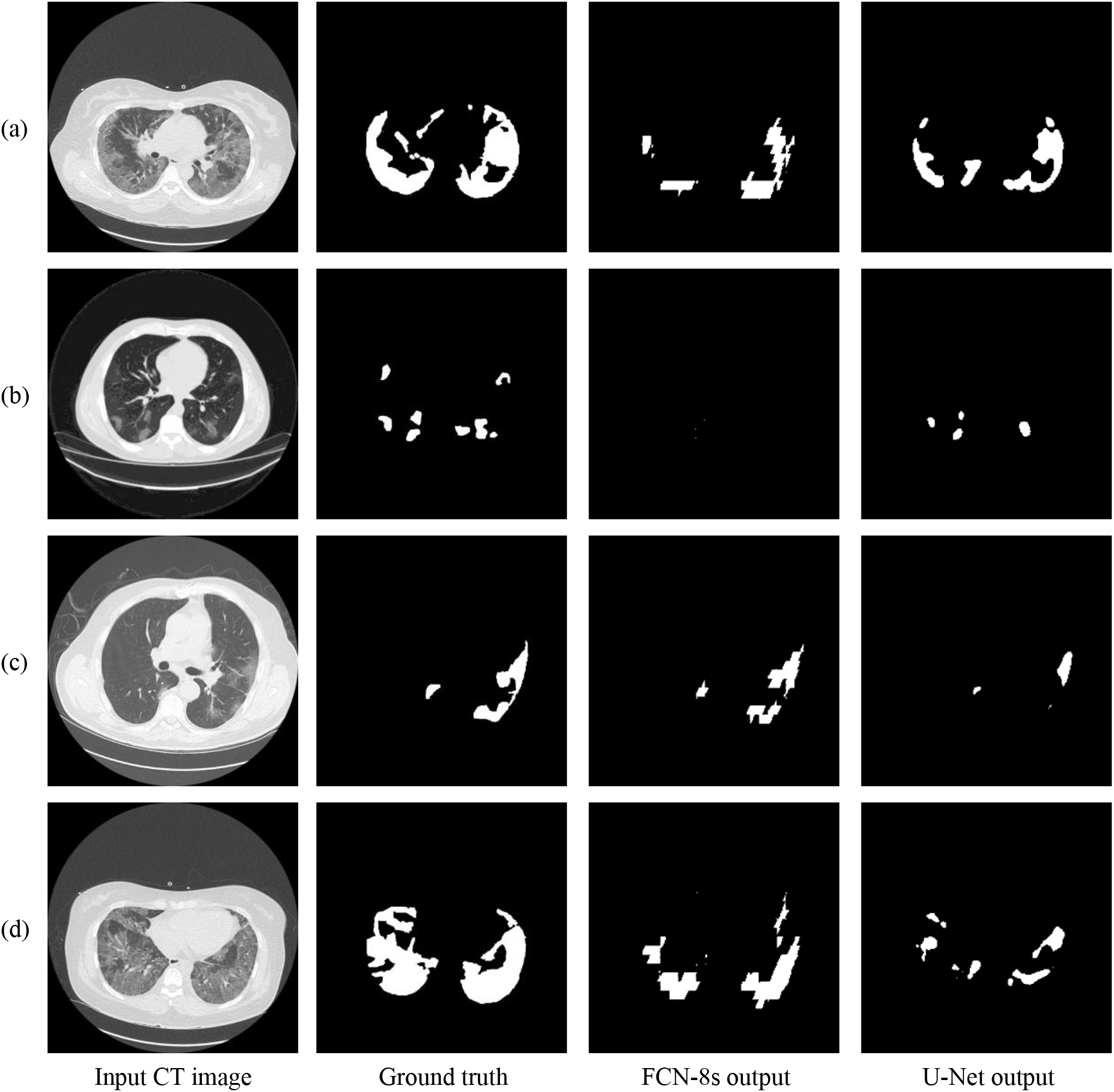
Relatively low detection performance on challenging images. More specifically, FCN-8s and U-Net show low detection accuracy in (a)-(b) and (c)-(d) respectively.

## IV. Conclusion

In this paper, we have presented a deep learning based approach for semantic segmentation in CT images for the detection of COVID-19 induced symptoms in the lung area. Preliminary results indicate that the proposed Fully Convolutional Neural Networks are capable of providing accurate segmentation for symptomatic areas, despite the class imbalance on the dataset and the man-made annotation errors on the boundaries of symptom manifestation areas, and could thus assist doctors in the detection as a fast and cost-effective supplementary option.

## Data Availability

The dataset used was collected from Radiopaedia.

https://radiopaedia.org/

